# AI-Assisted Pneumonia Detection, Localisation and Report Generation from Chest X-rays

**DOI:** 10.64898/2026.03.20.26348879

**Authors:** Federico E. Boiardi, Antoine D. Lain, Joram M. Posma

## Abstract

Pneumonia detection in chest X-rays (CXRs) is complicated by high inter-observer variability and overlapping radiographic patterns. While deep learning (DL) solutions show promise, limitations in generalisability and explainability hinder clinical adoption. We address these challenges by introducing a holistic DL-based computer-aided diagnosis (CAD) pipeline for pneumonia detection, localisation, and structured report generation from CXRs. We curated the largest composite of publicly available CXRs to date (N=922,634), of which 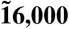 were used for training. MIMIC-CXR radiology reports were relabelled using a local large language model (LLM), positing that LLM-derived pneumonia labels would yield higher diagnostic sensitivity than the provided rule-based natural language processing (rNLP) labels. DenseNet-121 classifiers were trained on four configurations: MIMIC-CXR (rNLP), MIMIC-CXR (LLM), and each supplemented with VinDr-CXR data. Gradient-weighted Class Activation Mapping (Grad-CAM) provided visual explainability and lung zone-based localisation. LLM-driven relabelling significantly improved human-label agreement (96.5% vs 72.5%, P=1.66×10^−11^). The best-performing model (MIMIC-CXR (LLM) + VinDr-CXR) achieved 82.08% sensitivity and 81.97% precision, surpassing both radiologist sensitivity ranges (64-77.7%) and CheXNet’s pneumonia F1-score (43.5%). Grad-CAM localisation attained a moderate F1-score of 52.9% (sensitivity=65.7%, precision=44.3%), confirming focus alignment with pathological lung regions while highlighting areas for refinement. These findings demonstrate that LLM-driven label curation, combined with DL, can exceed conventional rNLP and radiologist performance, advancing high-quality data integration in predictive medical imaging. Clinically, our pipeline offers rapid triage, automated report drafting, and real-time pneumonia surveillance; tools that can streamline radiology workflows and mitigate diagnostic errors.

## Introduction

Pneumonia claims over 2 million lives annually despite being preventable and treatable (1). Diagnosis typically relies on radiological and pathological evidence, though treatment is often started based on the former alone (2). Chest X-rays (CXRs) are the primary imaging modality for evaluating suspected pneumonia given their speed, accessibility, and low cost (3). On CXRs, pneumonia manifests as white opacifications that contrast against the dark lung fields. These opacities reflect infiltrates such as consolidation, stemming from the influx of inflammatory cells and fluid into the alveoli.

However, high inter-observer variability in detecting pneumonia underscores the inherent subjectivity of radiographic interpretation (4, 5). This may result from the lower contrast of CXRs compared to other modalities, thereby being less effective at visualising soft tissue and distinguishing overlapping anatomical structures (6, 7). Time constraints exacerbate this challenge, with radiologists often spending just 5-10 seconds per image (8).

Given these limitations, computer-aided diagnosis (CAD) systems leveraging artificial intelligence (AI) have been developed to improve and automate CXR interpretation. Deep learning (DL) models trained on CXR data have achieved performance on par with experienced radiologists in detecting pneumonia and other pathologies (9–11), and 30-50% of radiologists already use or plan to use AI tools in their practice (12). Unfortunately, while many algorithms perform well in controlled studies, they often fail to generalise in real-world settings (13). One major culprit is reliance on noisy training labels, often derived from radiology reports using rule-based natural language processing (rNLP) (14). Models trained on such labels may inadvertently learn erroneous patterns that do not reflect genuine clinical findings, yielding deceptively high performance on internal datasets and poor generalisability in practice, especially when the model’s decision-making process lacks explainability (15), preventing clinical adoption.

This paper presents a DL-based CAD pipeline for pneumonia detection, localisation, and structured report generation from CXRs. Utilising a six-dataset composite of over 920K publicly available CXRs, we integrate radiology report relabelling and generation using large language models (LLMs) and employ Gradient-weighted Class Activation Mapping (Grad-CAM) to facilitate model explainability and localise opacities suggestive of pneumonia. We hypothesise that a model trained on a small set of human-verified labels, coupled with LLM-derived labels, will achieve a diagnostic sensitivity comparable to that of radiologists and outperform models trained on rNLP labels.

## Materials and Methods

### Data acquisition

Suitable CXR datasets were retrospectively identified through literature review and searches on PubMed, Google Scholar, and Kaggle using the terms “pneumonia dataset” and “chest X-ray dataset” (on 18 October 2024). Inclusion criteria were: public accessibility, provision of labelled CXRs with confirmed pneumonia diagnoses, and sufficient dataset size (over 5,000 images). Exclusion criteria included extensive poor image quality and restricted patient demographics. Of nine datasets initially identified, six met the criteria (Table 1). All datasets were downloaded programmatically.

**Table 1.** Publicly accessible chest X-ray (CXR) datasets suitable for pneumonia classification. Images/patients: number of images and patients in the dataset. Format: image file type. View: CXR projections (frontal or multi-view, with multi-view denoting both frontal and lateral images). Labels: number of distinct disease annotations per image (all include pneumonia). Reports: indicates whether images are accompanied by corresponding radiology reports (yes: Y, no: N). Demographic: indicates the country of origin of each dataset. Note: all patient data is already anonymised or pseudonymised in compliance with General Data Protection Regulation (GDPR). * Datasets requiring user credentialing or a signed data use agreement for full access.

| Dataset | Reference | Images | Patients | Format | View | Labels | Reports | Demographic |
| --- | --- | --- | --- | --- | --- | --- | --- | --- |
| MIMIC-CXR* | (16) | 377,110 | 65,379 | JPG | Multi | 14 | Y | USA |
| Stanford CheXpert* | (17) | 224,316 | 64,540 | JPG | Multi | 14 | N | USA |
| PadChest* | (18) | 160,861 | 67,625 | PNG | Multi | 19 | Y | Spain |
| ChestX-ray14 | (19) | 112,120 | 30,805 | PNG | Frontal | 15 | N | USA |
| RSNA Pneumonia | (20) | 30,227 | 26,684 | DICOM | Frontal | 2 | N | USA |
| VinDr-XR* | (21) | 18,000 | 18,000 | DICOM | Frontal | 28 | N | Vietnam |
| total |  | 922,634 | 273,033 |  |  |  |  |  |

We used subsets of the MIMIC-CXR (16) and VinDr-CXR (21) datasets for training, and evaluated model performance on independent data drawn from both. VinDr-CXR contains radiologist annotations, while MIMIC-CXR provides corresponding verbatim radiology reports for each image. We also evaluated models on four additional datasets: CheXpert (17), PadChest (18), ChestX-ray14 (19), and RSNA-Pneumonia (20), which are a mixture of expert (RSNA) and rNLP-generated labels (others). These data have been previously reported in isolation, but this manuscript represents the first time these have been combined.

The MIMIC-CXR data use agreement prohibits sharing images and reports. In compliance, all MIMIC-CXR images in this paper have been substituted with similar examples from datasets that permit public distribution, following established precedent (11). All X-ray images indicate the data source and, where relevant, the original dataset ID used for inference. Report snippets shown are synthetically generated using locally-deployed, open-weight LLMs, though they resemble the original MIMIC-CXR report text. IDs are disclosed in figure captions to allow those with data access to inspect the original images/reports.

### Image standardisation and view classification

DI-COM images were standardised using associated metadata. Grayscale inversion was corrected using Photometric Interpretation tags, and intensity ranges clipped according to Window Center and Width parameters. CXRs from PadChest and VinDr-CXR were downscaled to 8-bit resolution via minmax normalisation to preserve relative contrast. CXR projections were then classified as frontal or lateral using a residual network model previously trained on MIMIC-CXR data (11). Since pneumonia is predominantly diagnosed from frontal views, lateral CXRs were excluded from subsequent processing.

### Trunk segmentation, background clipping and contrast enhancement

CXRs were segmented to isolate the trunk through a pre-trained pyramid scene parsing network model from torchxrayvision (Fig. 1). Following this, background pixels were clipped to 0 intensity (i.e., pure black) and contrast was enhanced through Contrast Limited Adaptive Histogram Equalisation (22) (Fig. 2); see Supplementary Methods for additional details. Pre-processed images were exported as lossless PNG files.

**Fig. 1.**
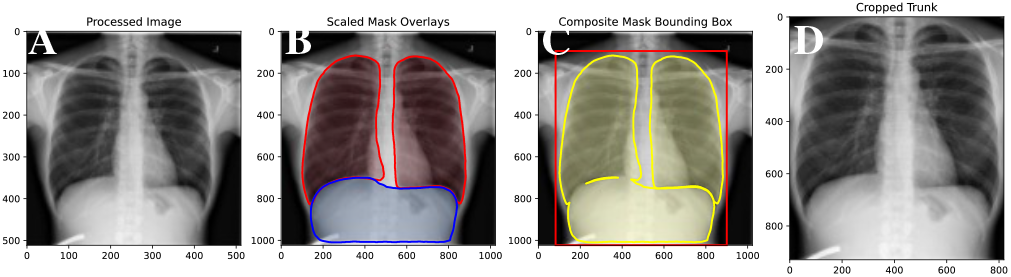
Automated cropping of trunk region from a chest X-ray (CXR). A: CXR after normalisation and resizing (512×512 pixels). B: Scaled masks of lungs (red) and abdomen (blue) overlaid on the original image (1024×1024 pixels). C: Composite mask (yellow) with bounding box (red) (+2.5% padding). D: Cropped CXR focusing on the trunk. CXR source: ChestX-ray14 (19).

**Fig. 2.**
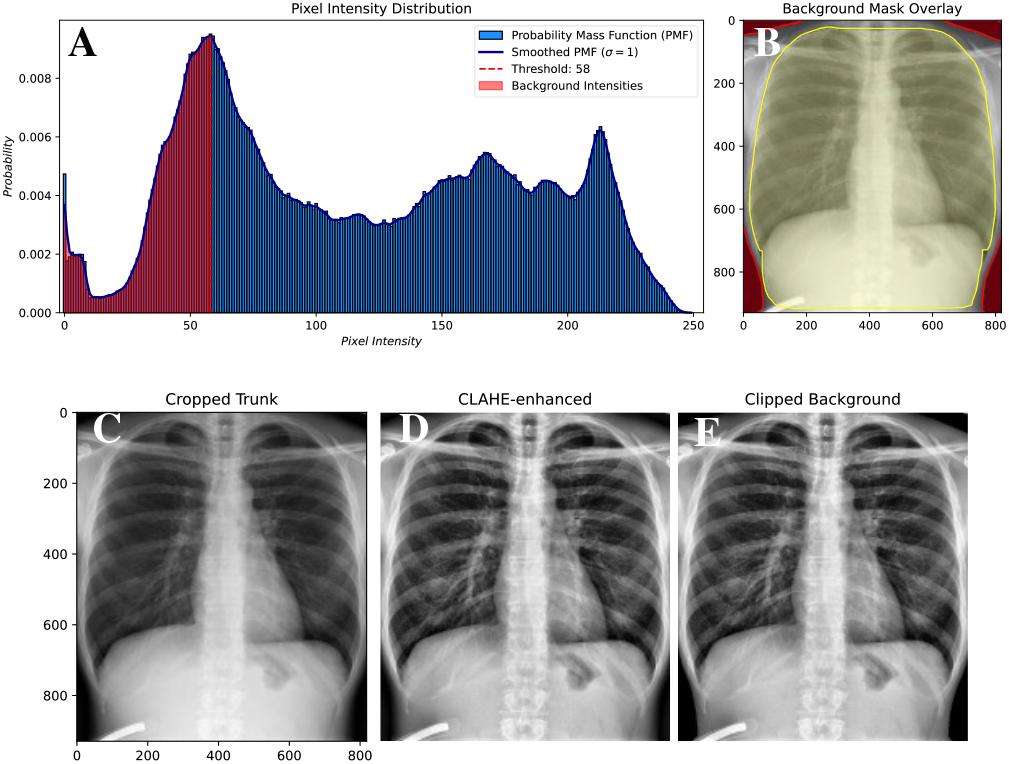
Background clipping and contrast enhancement of a chest X-ray (CXR). A: Cropped CXR pixel intensity probability mass function. B: Cropped CXR showing background mask overlay (red) outside the filled composite mask (yellow). C: Cropped CXR. D: CXR after applying Contrast Limited Adaptive Histogram Equalisation (CLAHE). E: CLAHE-enhanced CXR with clipped background. CXR source: ChestX-ray14 (19).

### Quality control

Poor-quality CXRs were flagged during view classification (Fig. 3A-C) and exception handling within the pre-processing pipeline (Fig. 3D-F). Discrepancies between reported and predicted CXR projections often reflected poor quality, with distortions that obscured or eliminated relevant diagnostic details. Images were also excluded from downstream analysis if either segmented lung mask was absent or the composite mask contained more than three connected components (i.e., non-anatomical artefacts).

**Fig. 3.**
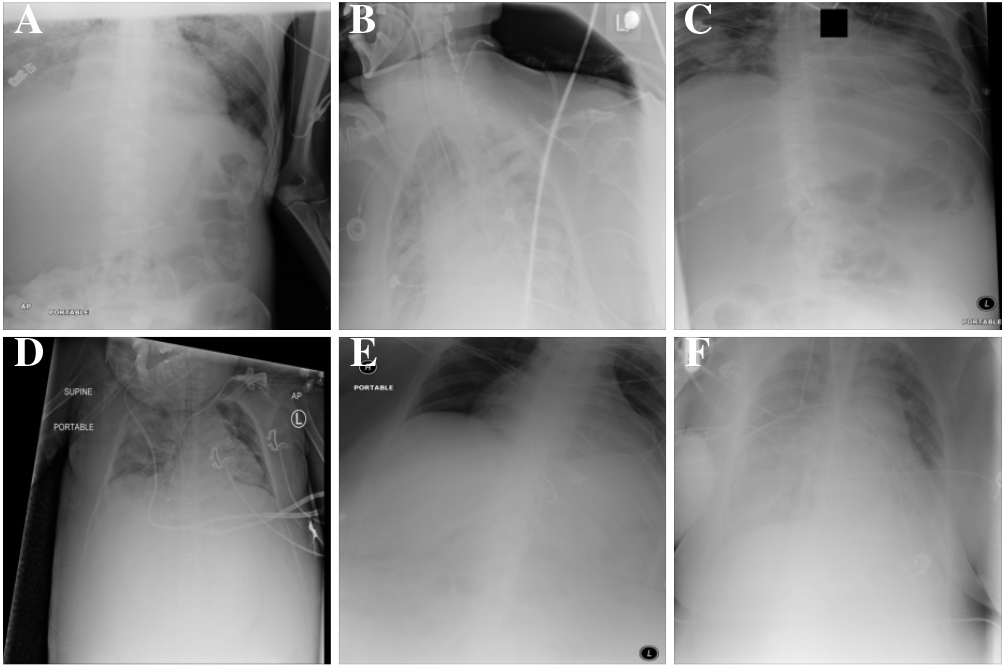
Sample of identified poor-quality chest X-rays (CXRs). A-C: Examples identified during view classification. D-F: Examples where the segmentation model failed to accurately locate the lungs. CXR source: ChestX-ray14 (19).

### Label extraction

Most datasets from Table 1, including MIMIC-CXR, contain labels extracted using rNLP methods, such as those developed from the CheXpert dataset. During preliminary inspection of MIMIC-CXR metadata, >200 pneumonia-positive labels were identified despite corresponding radiology reports explicitly stating “no findings to suggest pneumonia”, or equivalent negations. To mitigate the aforementioned generalisability issues associated with noisy labels, MIMIC-CXR reports were relabelled using a local LLM. The findings, impressions, and conclusions sections (where available, otherwise substituted by other sections) from 214,332 reports were used to identify reports with any mention of pneumonia (e.g. case-insensitive strings like “pneumonia” or “infectio”; case-sensitive abbreviations such as “HAP” (hospital-acquired pneumonia)). Relevant reports were passed to a locally-deployed reasoning LLM (`DeepSeek-R1-Distill-Llama-8B`, temperature=0.6, constant randomState) with a custom prompt (see Supplementary Methods, Fig. 7) to classify reports as pneumonia-positive, -negative, or -uncertain, and to extract positional descriptors if present. These revised labels were used for model training along with VinDr-CXR data, where the majority vote was adopted as the final label. The remaining datasets were excluded from training due to the lack of accompanying full radiology reports needed to verify label accuracy.

### Label verification

A random sample of 200 MIMIC-CXR reports, mirroring the original label distribution, was independently reviewed by three annotators to extract pneumonia labels. Disagreements were resolved to obtain a single set of agreed-upon human labels. Human agreement with original rNLP and LLM labels was measured according to strict agreement and Cohen’s kappa coefficient (*κ*). A one-sided, two-proportion Z-test was used to determine if LLM labels had significantly higher agreement with human labels compared to rNLP labels, with *P*<0.01 indicating rejection of the null hypothesis.

### Deep learning model architecture and training

Classification models were trained using four distinct dataset configurations. Two baseline models were trained exclusively on MIMIC-CXR data (18,354 images) with either rNLP or LLM labels. Two composite models were trained by integrating VinDr-CXR with the respective MIMIC data (19,764 images). Data was split into training, validation, and test sets (80:10:10), ensuring balanced class representation and no patient ID leakage. Training sets were augmented through a series of stochastic transformations (horizontal flipping, affine rotations (×5°) and translations (±5%), brightness/sharpness adjustments, Gaussian blurring).

A DenseNet-121 architecture with ImageNet pre-trained weights (23) was initialised with a custom classification head (fully connected 512-unit layer, ReLU activation, batch normalisation, 30% dropout), outputting two logits (pneumonia-positive vs. -negative). Images were resized to 480 480 pixels and normalised using ImageNet channel means and standard deviations.

Training was performed with GPU-acceleration, batch size 32, and 4 parallel workers for approximately 50 epochs. Cross-entropy loss was minimised using the Adam optimiser (learning rate 5×10^−5^, weight decay 1×10^−3^). Learning rate was adjusted based on validation loss (ReduceLROnPlateau, factor=0.1, patience=3 epochs). Early stopping was invoked after 5 consecutive epochs without sensitivity improvements, and the model achieving the highest validation sensitivity was saved. All reported thresholds and hyperparameters were tuned via grid-search optimisation on validation data.

### Deep learning model evaluation and error analysis

Model performance was assessed on the MIMIC (LLM) + VinDr-CXR test set according to precision, sensitivity, F1-score, and area-under-the-receiver-operating-characteristic curve (AUC). The best-performing model was evaluated on test sets derived from all six datasets (16–21), each proportionally sampled based on the number of pneumonia-positive images in each dataset with performance measured across nine independent runs using different random states.

MIMIC-CXR test set CXRs were stratified into four groups: positive, negative, uncertain, and N/A (no mention of pneumonia). The model was applied to each CXR to obtain pneumonia probability scores (0-100%). For each group, kernel density estimates were computed over the predicted probabilities, with pairwise comparisons done using the Kruskal-Wallis test, followed by Bonferroni-corrected Dunn’s posthoc test.

A targeted error analysis was conducted on false-negative reports (N=188) to uncover linguistic patterns associated with model misclassification (see Supplementary Methods).

### Deep learning model explainability

Grad-CAM (24) heatmaps were thresholded using Otsu’s method (25) and mapped onto anatomically defined lung zones to predict pneumonia localisation. Pneumonia-positive CXRs from the MIMIC-CXR test set were processed by segmenting the lung masks into three zones of equal height (upper, middle, lower) and considered pneumonia-positive if the maximum Grad-CAM activation exceeded 0.8 (i.e., 80% of the global maximum). Localisation performance was evaluated by comparing reported and predicted pneumonia-positive lung zones according to precision, sensitivity, and F1-score. Report text was generated using the predicted localisation using `DeepSeek-R1-Distill-Llama-8B`. See Supplementary Methods for further details.

### Software

All computations and visualisations were implemented in Python v3.9.21 using the `nltk` (v3.9.1), `opencv` (v4.10.0.84), `pytorch` (v2.5.1), `scikit-learn` (v0.11.4), `scipy` (v1.13.1), `statsmodels` (v0.14.4), `torchxrayvision` (v1.3.2), and `transformers` (v4.48.1) packages on a Linux workstation (Ubuntu 6.8.0) with two NVIDIA RTX4090 GPUs (2×24GB). Source code and prompts are available on GitHub (url-removed-for-anonymisation).

## Results

### Relabelling evaluation

Several open-weight LLMs were evaluated for relabelling, with `DeepSeek-R1-Distill-Llama-8B` showing the best performance with 96.5% agreement with manually extracted labels (Table 2).

**Table 2.** Performance of open-weight large language models (LLMs) on re-classifying pneumonia labels based on reports. Agreement given for the independent, manually labelled set of 200 reports.

| Model | Agreement |
| --- | --- |
| DeepSeek-R1-Distill-Llama-8B | 96.5% |
| Mistral-7B-Instruct-v0.2 | 93.5% |
| Qwen2.5-3B-Instruct | 88.5% |
| Llama-3.2-3B-Instruct | 87.0% |
| gemma-3-1b-it | 81.5% |

We compare the pneumonia label distributions in MIMIC-CXR under the original rNLP pipeline versus LLM-based relabelling in Table 3. LLM relabelling roughly halved positive cases (−46.85%), and moderately increased negative (+39.91%) and uncertain cases (+25.06%). There was a marginal reduction in N/A labels (−4.55%). From the sample of human-annotated reports, agreement was moderate with rNLP labels (*κ*=0.49) and near-perfect with LLM labels (*κ*=0.93). Moreover, strict agreement was significantly higher with LLM labels (96.50%) compared to rNLP labels (72.50%) (*P*=1.66×10^−11^).

**Table 3.** MIMIC-CXR pneumonia label counts for original rule-based natural language processing (rNLP) and new large language model (LLM) methods. * N/A: No pneumonia label provided/available, with new labels determined by rule-based absence of pneumonia-related terms rather than by an LLM.

| Pneumonia label type | rNLP labels | LLM labels |
| --- | --- | --- |
| Positive | 26,222 | 13,937 |
| Negative | 43,358 | 60,664 |
| Uncertain | 29,150 | 36,455 |
| N/A | 278,365 | 265,692* |

### Pneumonia classification and error analysis

Models trained on LLM labels demonstrated an average improvement of 4-5% in precision and sensitivity relative to their rNLP counterparts (Fig. 4A-B). Incorporating VinDr-CXR training data further improved performance by 0.5-2%. The best-performing model, trained on the combined MIMIC (LLM) and VinDr-CXR data, achieved a mean sensitivity of 82.08% and precision of 81.97% for pneumonia-positive cases.

**Fig. 4.**
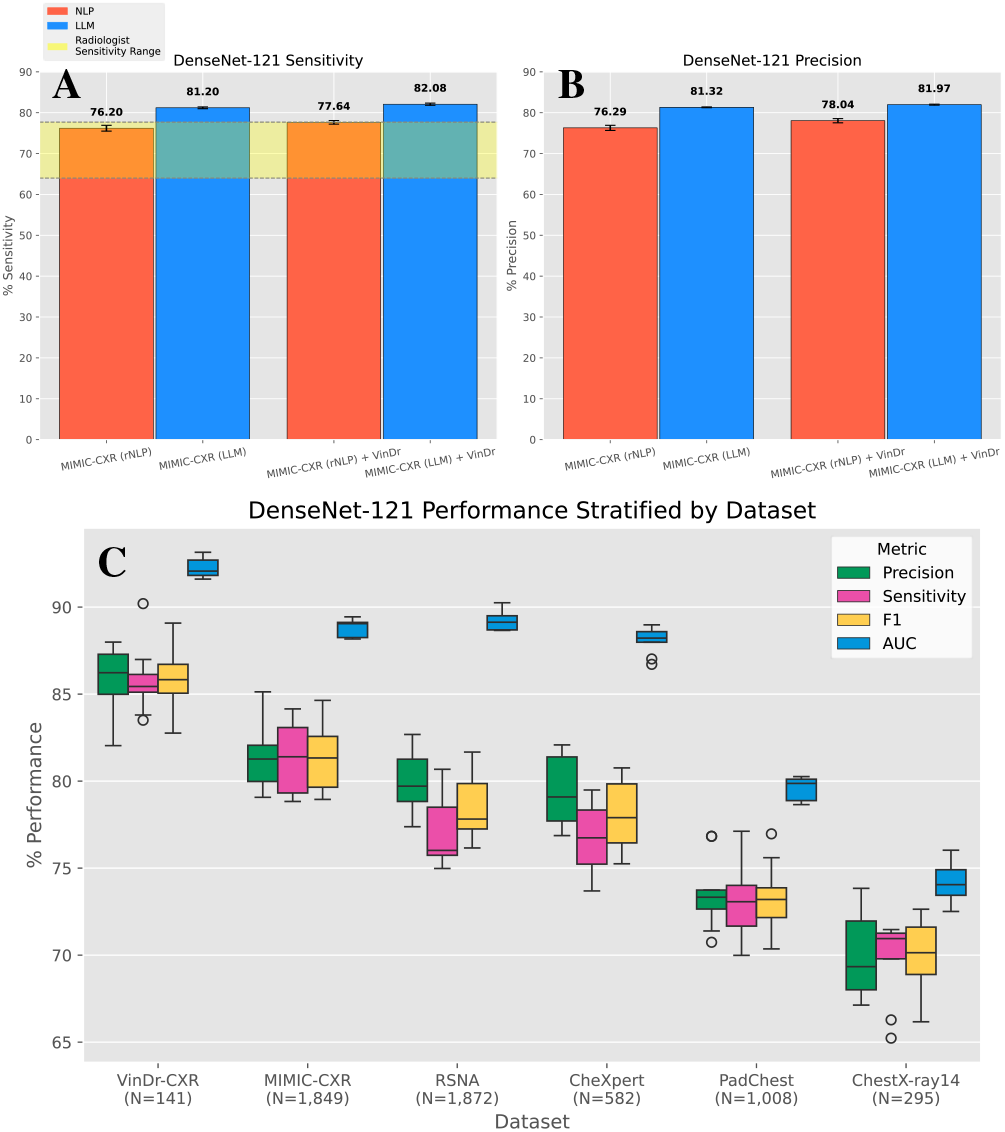
Test set sensitivity (A) and precision (B) of DenseNet-121 models in detecting pneumonia, trained using four distinct sets of labels: MIMIC (rNLP), MIMIC (LLM), MIMIC (rNLP) + VinDr-CXR, and MIMIC (LLM) + VinDr-CXR. Results are presented as mean × standard deviation over 9 test runs. The shaded yellow region in panel A denotes the reported radiologist sensitivity range for pneumonia detection in chest X-rays. C: Boxplot comparison of MIMIC LLM + VinDr-CXR DenseNet-121 performance metrics for pneumonia detection across six chest X-ray (CXR) datasets. Each boxplot represents the distribution of 9 test runs. AUC: Area Under the Curve.

When evaluated across all datasets (Table 1), VinDr-CXR had the highest average test set performance, with an F1-score of 85.83% and AUC of 92.23% (Fig. 4C). MIMIC-CXR followed closely, achieving an average F1-score of 81.35% and AUC of 88.78%. RSNA and CheXpert demonstrated comparable performance, each obtaining an approximate F1-score of 78% and AUC between 88-89%. PadChest exhibited moderately lower performance (F1=73.33%, AUC=79.52%), while ChestX-ray14 showed the lowest performance overall (F1=69.85%, AUC=74.18%).

Figure 5 delineates the model’s predicted pneumonia probability distributions for each ground-truth label category in the MIMIC-CXR (LLM) test set. Pneumonia-positive images were associated with high confidence scores (median=89.53%), while negative images clustered at the low end (median=18.21%). Uncertain cases showed a broader distribution, skewed toward higher probabilities (median=71.91%, interquartile range (IQR)=53.20%). The N/A group showed the broadest distribution (IQR=54.90%), appearing bimodal though primarily spanning mid-to-low probabilities (median=30.13%). Overall, the model was significantly more confident in the presence of pneumonia among pneumonia-positive images compared to negative ones (Kruskal-Wallis and Dunn’s tests, *P*<1×10^−16^).

**Fig. 5.**
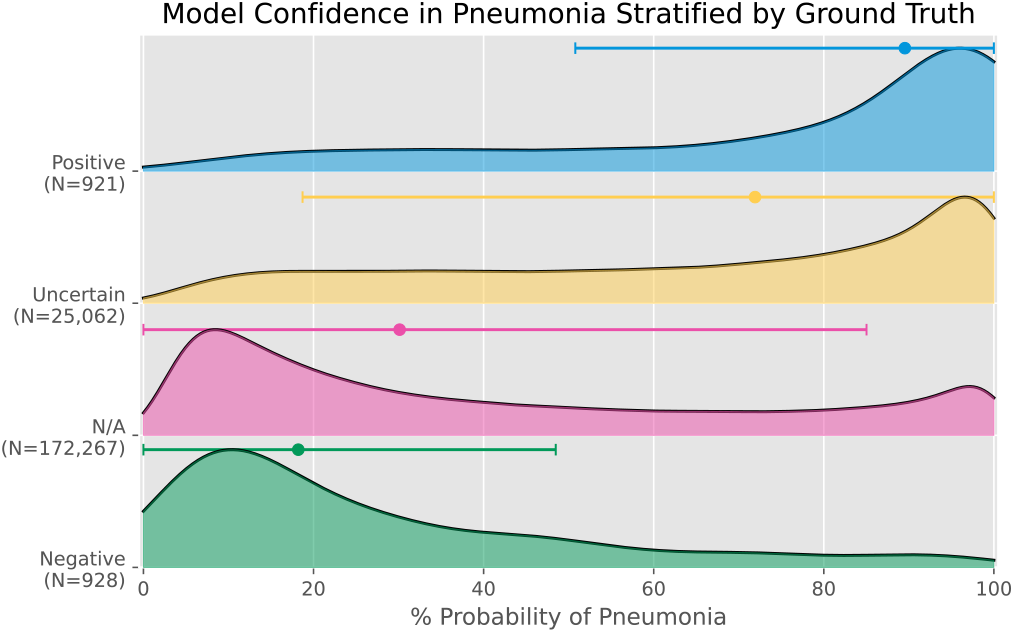
Ridgeline plot of predicted pneumonia probability distributions for each ground-truth label category in the MIMIC-CXR (LLM) test set. Each group is represented by its Gaussian kernel density estimate, with error bars denoting median ± interquartile range (IQR). N/A: No mention of pneumonia in radiology reports.

N-gram analysis of 188 false-negative reports highlighted frequently recurring descriptors such as “left lower” (N=92), “left lower lobe” (N=76), and “left lower lobe pneumonia” (N=24). These phrases ranked among the top-ten n-grams by mean TF-IDF score within false-negative reports, indicating their overrepresentation in these misclassified cases.

### Pneumonia localisation

Visual inspection of Grad-CAM heatmaps from pneumonia-positive predictions confirmed that the model’s attention is exclusively focused on the lung fields, often coinciding with radiologic signs associated with pneumonia (e.g., consolidation) (Fig. 6). Quantitative comparison of heatmap activations and radiologist annotations yielded a localisation F1-score of 52.89% on the MIMIC-CXR test set (sensitivity=65.65%, precision=44.29%).

**Fig. 6.**
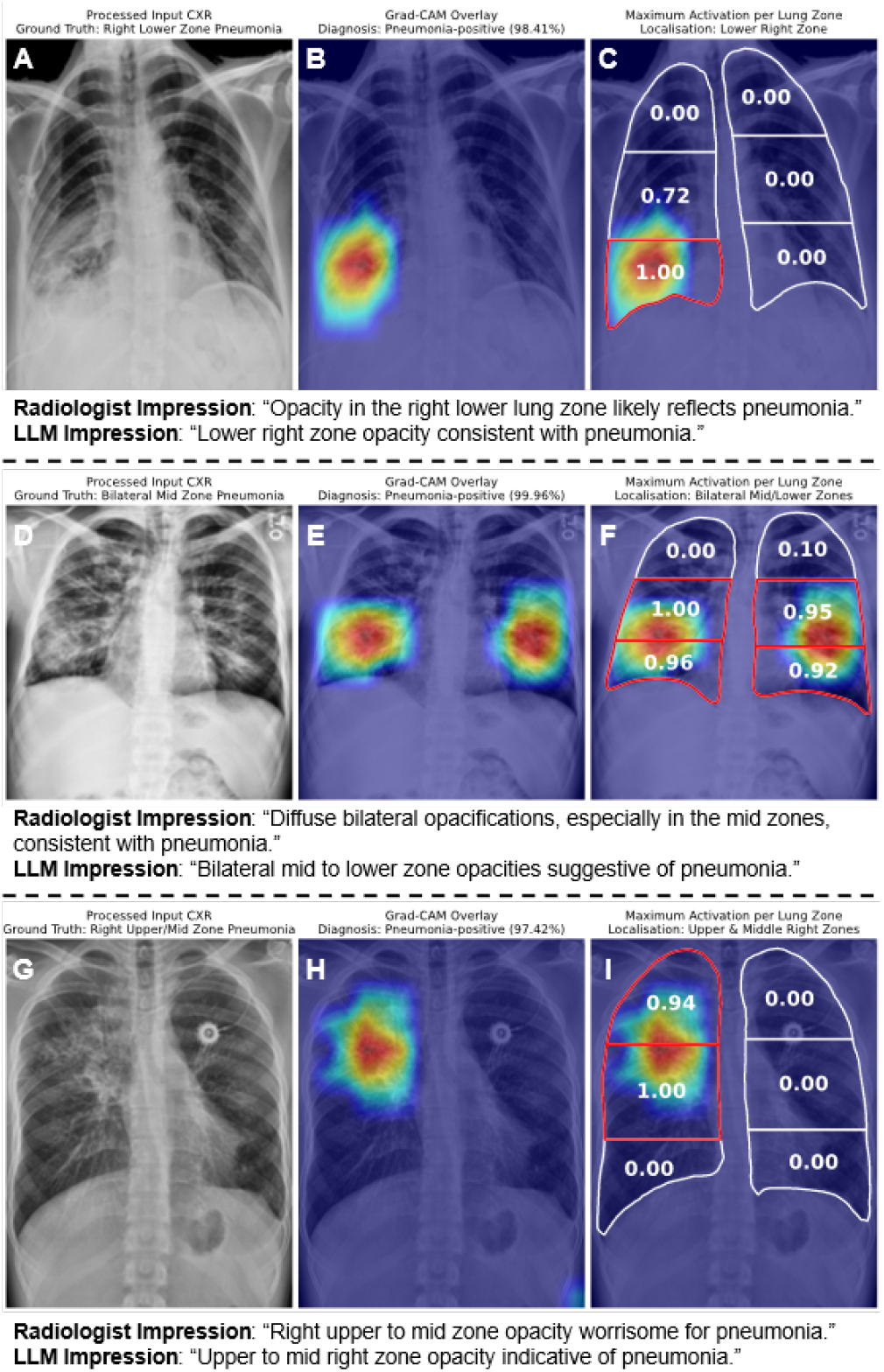
Pneumonia localisation in chest X-rays (CXRs) using Gradient-weighted Class Activation Mapping (Grad-CAM). A: CXR showing pneumonia-associated opacification in the right lower zone (left side of the image). ChestX-ray14 ID: 00000261_002; Original MIMIC-CXR ID: 52aa2f0a-932bb134-bb4b4d53-ca4fb145-9d6966b5. B: Grad-CAM heatmap for the pneumonia-positive class (98.41% confidence) superimposed on the CXR. Heatmap is thresholded using Otsu’s method to isolate higher activations (red), which localise to the region of opacification. C: Segmented lung fields are partitioned into three zones (upper, middle, and lower), with each displaying its maximum activation value. Heatmap is masked to exclude non-lung regions and renormalised to the [0,1] range. Zones exceeding 80% of the global maximum (i.e., 0.8) are classified as pneumonia-positive (indicated by red contours). In this example, the model’s attention correctly aligns with the pneumonia-positive right lower zone. D: Patient with diffuse pneumonia, primarily localised to the mid zones bilaterally. ChestX-ray14 ID: 00000963_007; MIMIC-CXR ID: 6043b47b-bfb45ef3-6ed9cb92-2381888f-9e91f2b8. E: Pneumonia-positive Grad-CAM heatmap (99.96% confidence) localised to the regions of opacification. F: Correct classifications of the bilateral mid zones, as well as the lower zones. G: Patient with pneumonia localised across the right upper and mid zones. ChestX-ray14 ID: 00001637_001; MIMIC-CXR ID: a46507be-27a0330c-dac7aeae-cbfab7d2-6829f9d1. H: Pneumonia-positive Grad-CAM heatmap (97.42% confidence) localised to the region of opacification. I: Correct classifications of the pneumonia-positive zones. CXR source: ChestX-ray14 (19).

## Discussion

We presented a pipeline to aggregate and pre-process publicly available CXR datasets (N=922,634) and trained a DL model capable of detecting pneumonia, achieving F1-scores of 69.85% to 85.82% depending on dataset and label quality. The three datasets with highest performance either had human-verified labels (VinDr-CXR, RSNA-Pneumonia) or full-text reports to permit LLM-based relabelling of rNLP labels (MIMIC-CXR). Finally, we demonstrated model explainability, both visually (Grad-CAM heatmaps) and textually (LLM-generated summaries).

We leveraged `DeepSeek-R1-Distill-Llama-8B` to relabel MIMIC-CXR reports due to its strength in reasoning compared to closed-source models (26). We focused on open-weight models to ensure our pipeline could be deployed locally and fine-tuned on proprietary data in hospital trusts, with flexible adaptation to other diseases. LLM-derived pneumonia labels showed strong agreement with human labels and markedly shifted class distributions; notably, halving positive cases in MIMIC-CXR. This suggests that the CheXpert labeller, used to label MIMIC-CXR, is overconfident in identifying pneumonia-positive mentions. Our findings mirror recent successes of GPT-based labellers, which also demonstrated significant improvements over CheXpert labels in MIMIC-CXR (27).

Training DL models on high-quality labels (human+LLM) yielded consistent 4-5% gains in precision and sensitivity relative to rNLP labels. DenseNet-121 was selected from a pool of architectures (including ResNet, EfficientNet, ViT) based on validation performance. Our best model (82.08%) exceeds reported radiologist sensitivities for CXR pneumonia detection (64-77.7%) (28, 29). The modest performance boost from incorporating VinDr-CXR (0.5-2%) demonstrates the value of diverse training data in boosting model robustness. The poorest performance was recorded for ChestX-ray14 (19), a widely-used dataset with documented label noise (14). We purposely excluded ChestX-ray14 from training, since DL models trained on it have shown poor generalisability despite high internal accuracy (30). Pneumonia probability outputs aligned with expectations: true-positive and true-negative distributions were well-separated, while N/A and uncertain cases fell into the mid-range, reflecting genuine diagnostic ambiguity. False negatives were disproportionately associated with lower left pneumonia. This coincides with known radiological challenges: the heart and cardiac silhouette overlap with the lower left lung in frontal scans and may subsequently obscure opacifications (31). In retrospect, inclusion of lateral CXRs may have helped the model resolve such cases. Intriguingly, some N/A images, whose reports do not mention pneumonia, received high probability scores, suggesting potential missed cases that could benefit from AI-assisted triage. These probability scores can serve as risk estimates and be calibrated to suit different clinical thresholds.

Our model is explainable, with Grad-CAM activations largely confined to the lungs and often overlapping with infiltrates for pneumonia-positive cases. The model reliably identifies most pneumonia-positive zones (moderate sensitivity=65.65%), but a substantial proportion of activated regions extend beyond these zones (precision=44.29%), reflecting the lower bound of Grad-CAM localisation performance. For instance, if a report specifies ‘upper right zone’ but the model identifies both upper and middle right zones, inclusion of the middle zone is penalised (despite being anatomically adjacent), even if it is one continuous activation region. Different scoring functions may give more realistic estimates, though Grad-CAM visualisations nonetheless affirm our model’s capacity to focus on clinically relevant areas. While Grad-CAM has previously been used to visualise pneumonia classification, our method is constrained to the lung area and does not show spurious activations outside the thorax unlike prior work (32, 33).

### Study and Future Work

Our study has several limitations. Despite aggregating nearly one million images, only a small subset (16,000) was used for training; considerably fewer than the 100,000 used to train CheXNet on ChestX-ray14 (9). This reduction was due to quality filtering (e.g., poor-quality images, lateral views), absent pneumonia labels or report indications, and LLM-driven relabelling of MIMIC-CXR. Pneumonia-positive CXRs represented a small fraction of all images as most of the datasets, except RSNA-Pneumonia, are designed for multi-label pathology classification. The reduced training size was partially remedied using transfer learning and data augmentation. Of the datasets included, only MIMIC-CXR contained full reports suitable for LLM-based relabelling. PadChest provides stemmed reports in Spanish (18), but relabelling accuracy on these was poor, so rNLP labels were retained. In principle, our approach is applicable to any dataset with English reports and could be extended as such data becomes available. While the test sets differ, our model still outperforms CheXNet, with the added benefit of free-text report generation. Training was limited to two datasets with high-quality labels to allow for more general evaluation; for this reason RSNA-Pneumonia was reserved exclusively for testing as a human-labelled benchmark. Additional training data could have further improved our model at the cost of losing independent evaluation data. Another approach considered, but not explored, was to adjust learning rates based on label confidence (i.e., lower for rule-based labels, and higher for human-labelled data).

Our reliance on public datasets inherently introduces demographic biases, and there is no guarantee that the model will generalise across all clinical settings. Likewise, comparisons with radiologist performance are based on published benchmarks as opposed to a unified reader study. However, one of our test datasets is fully human annotated and can serve as a more realistic benchmark. Generated report text was based on the model’s classification and Grad-CAM localisation. Due to the apparent improvements that can be made to the location detection, we did not assess the accuracy of these image-sentence pairs through independent radiologist review, as done in prior work (11). Future efforts should include this validation step, alongside extension of the pipeline to multilabel thoracic pathology classification, evaluation across diverse populations (e.g., paediatric datasets), and prospective validation in clinical environments.

In summary, we developed a pneumonia detection pipeline with performance comparable to that of radiologists, capable of explaining decision making, and generating free-text reports. Our work, demonstrated on six datasets, underscores the transformative potential of high-quality data and the versatility of AI techniques in medical imaging. In high-volume or resource-limited settings, an automated assistant that consistently achieves human-level sensitivity could reduce diagnostic delays, minimise oversight errors, and free specialists to prioritise complex cases, ultimately improving patient out-comes.

## Data Availability

All data used are available through credentialised access at: https://physionet.org/content/mimic-cxr/2.1.0/, https://stanfordmlgroup.github.io/competitions/chexpert/, https://bimcv.cipf.es/bimcv-projects/padchest/, https://nihcc.app.box.com/v/ChestXray-NIHCC, https://www.rsna.org/artificial-intelligence/ai-image-challenge/rsna-pneumonia-detection-challenge-2018, https://vindr.ai/cxr.

https://stanfordmlgroup.github.io/competitions/chexpert/

https://bimcv.cipf.es/bimcv-projects/padchest/

https://nihcc.app.box.com/v/ChestXray-NIHCC

https://www.rsna.org/artificial-intelligence/ai-image-challenge/rsna-pneumonia-detection-challenge-2018

https://vindr.ai/cxr

## AUTHOR CONTRIBUTIONS

Conceptualisation, J.M.P. and A.D.L.; Methodology, F.E.B., A.D.L. and J.M.P.; Formal Analysis and Investigation, F.E.B.; Supervision, J.M.P., and A.D.L.; Writing – Original Draft: F.E.B., A.D.L. and J.M.P.; Writing – Review & Editing: J.M.P., and A.D.L. All authors have read and agreed to the published version of the manuscript.

## Acknowledgement

The NIH Clinical Center was the provider of the de-identified images of chest x-rays shown in this manuscript, and is available through the NIH download site: https://nihcc.app.box.com/v/ChestXray-NIHCC. All data used is publicly available with credentialised access and subject to data use agreements. Requests for data should be directed to the provider.

## Supplementary Materials

### Supplementary Methods

#### Trunk segmentation, background clipping and contrast enhancement

CXRs were cropped to isolate the trunk and remove diagnostically irrelevant features (e.g., text overlays). Trunk segmentation was performed using a pre-trained pyramid scene parsing network model from torchxrayvision (https://github.com/mlmed/torchxrayvision). CXRs were normalised to an intensity range of [-1024,1024] and resized to 512×512 pixels to match the model’s input requirements (Fig. 1A). Probability maps of the lungs and abdomen were derived from the model’s output logits via sigmoid activation. These maps were then rescaled to the original image dimensions using bilinear interpolation and thresholded at 50% to generate binary masks (Fig. 1B). The masks were combined into a composite mask, around which a bounding box was computed to define the cropping area (Fig. 1C), resulting in a focused view of the trunk (Fig. 1D).

Background pixels were identified based on the intensity distributions of the cropped CXRs. A probability mass function (PMF) of pixel intensities was calculated and smoothed using a Gaussian filter to reduce noise (Fig. 2A). The lowest local maximum in the smoothed PMF was selected as the background intensity threshold. To preserve diagnostically relevant regions, gaps between lung and abdomen masks were filled to create a continuous mask (Fig. 2B). This was achieved by horizontally and vertically connecting the outermost non-zero pixels within the mask’s bounding box. Pixels outside the continuous mask with intensity values below the threshold were classified as background (Fig. 2B). Only background-connected components touching the image borders were retained as true background to preserve dark anatomical regions (e.g., costophrenic angles). Contrast Limited Adaptive Histogram Equalisation (22) (CLAHE, clip limit=2) was applied to enhance local contrast using OpenCV (Fig. 2D). Following CLAHE, background pixels were clipped to an intensity of zero if they exceeded 5% of the image area to avoid unnecessarily altering images with minimal background noise (Fig. 2E). Finally, the pre-processed images were exported as lossless PNG files.

#### Label Extraction Prompt

Pneumonia labels were extracted from relevant reports using `DeepSeek-R1-Distill-Llama-8B` (https://huggingface.co/deepseek-ai/DeepSeek-R1-Distill-Llama-8B) (temperature=0.6), a locally-deployed LLM with reasoning capabilities. A custom prompt was designed to emulate an experienced radiologist’s analysis (below), instructing the model to classify reports as pneumonia-positive, pneumonia-negative, or uncertain for pneumonia. In addition, positional descriptors, when present, were extracted. Responses followed an XML tagging format, which were parsed using regular expressions to derive the final labels. A single random state was maintained across all queries. The prompt (‘chat’) in Fig. 7 was used as input to the locally-deployed `DeepSeek-R1-Distill-Llama-8B` model, with the code in Fig. 8 used to extract the relevant responses for each report appended to the instructions.

**Fig. 7.**
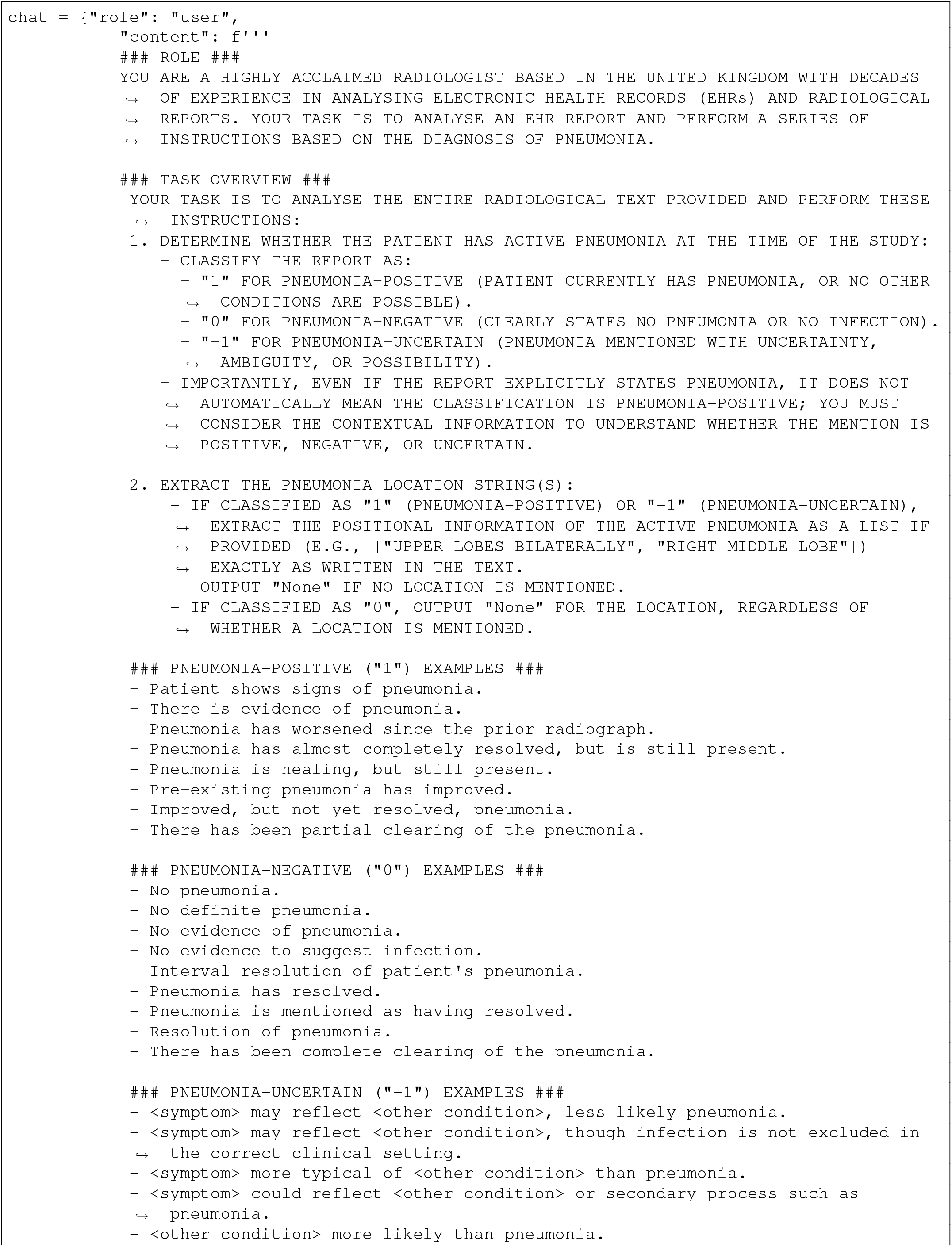
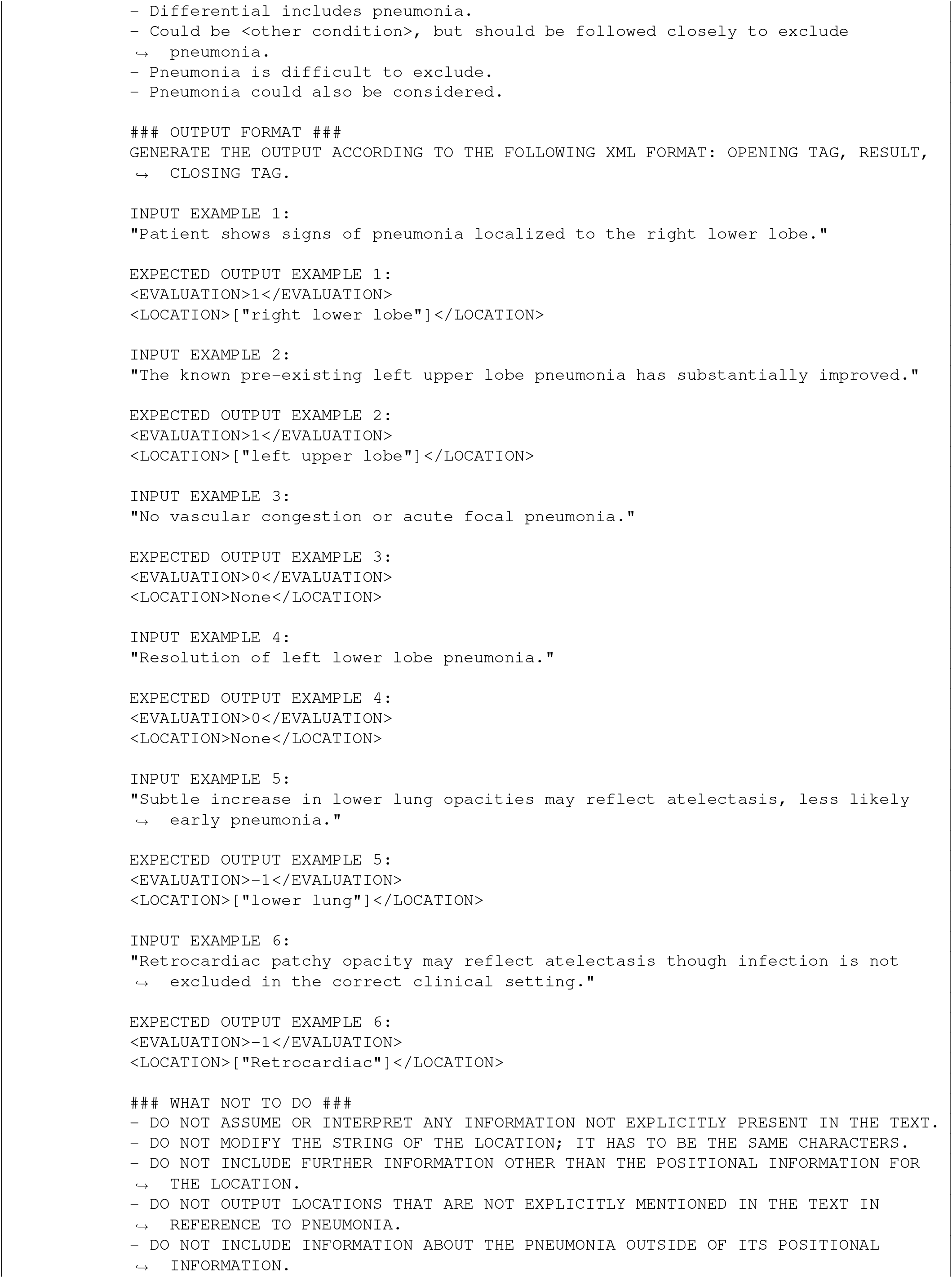
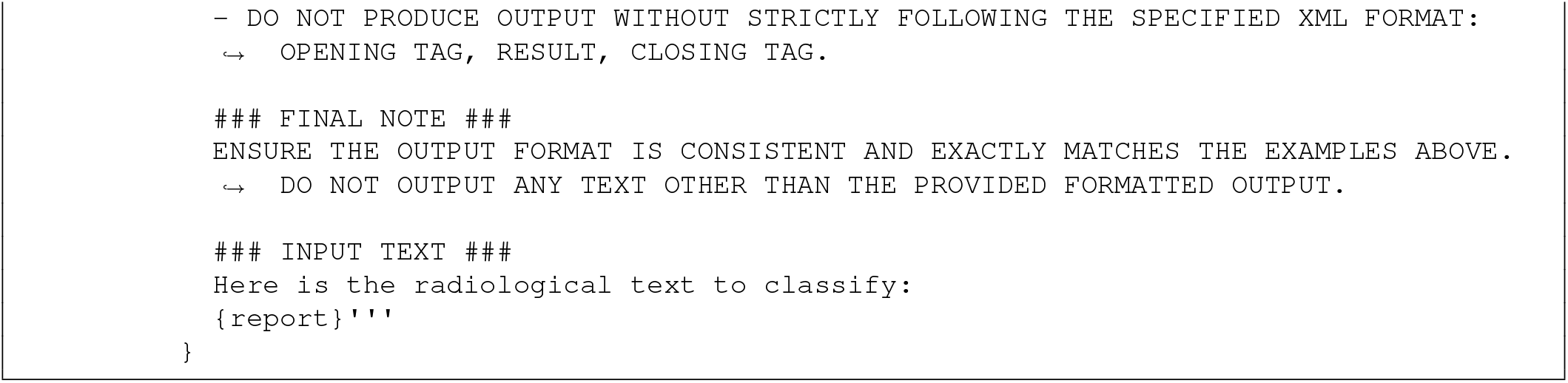
Prompt used as input to the locally-deployed DeepSeek-R1-Distill-Llama-8B model.

**Fig. 8.**
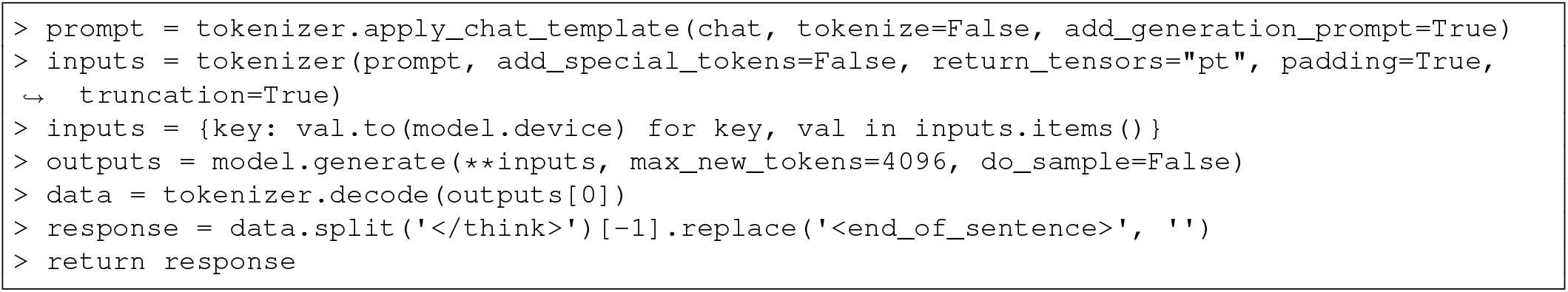
Code used to run the prompt.

#### Deep learning model error analysis

Reports were pre-processed through lowercasing, punctuation removal, to-kenisation, stop-word removal, and lemmatisation using nltk. Common phrases were extracted via n-gram analysis (n=2,3,4) and ranked by term frequency. A term frequency-inverse document frequency (TF-IDF) analysis was performed on the combined corpus of false-negative and true-positive reports using scikit-learn. Mean TF-IDF scores were computed for each group, and their differences were used to identify phrases disproportionately characteristic of false negatives.

#### Deep learning model explainability (extended)

Gradient-weighted Class Activation Mapping (Grad-CAM) (24) was implemented to interpret the model’s decision-making process. The approach captures activations from the final convolutional layer during the forward pass and gradients during backpropagation. To circumvent DenseNet’s in-place ReLU operations, the model’s forward function was patched to use a non-in-place variant, ensuring that hooked tensors remain unaltered. During inference, class probabilities are computed using a softmax function, and the predicted class is selected. A one-hot encoded gradient is then backpropagated to re-trieve class-specific gradients, which are globally averaged across spatial dimensions to yield importance weights. These weights are combined with the stored activations to generate a coarse heatmap that is subsequently rectified (via ReLU) and normalised to [0,1]. When superimposed on the original CXR, the heatmap highlights anatomical regions that influence the model’s diagnosis, providing qualitative insight into whether its focus aligns with clinically relevant features.

To predict pneumonia localisation, Grad-CAM heatmaps were mapped onto anatomically defined lung zones. Pneumonia-positive CXRs from the MIMIC-CXR test set were processed by segmenting the lung masks into three zones of equal height (upper, middle, and lower). The LLM-derived pneumonia locations were then assigned to the corresponding zones. For each true-positive prediction, the resulting Grad-CAM heatmap was thresholded using Otsu’s method (25), masked to exclude non-lung regions, and renormalised to [0,1]. A lung zone was classified as pneumonia-positive if its maximum activation exceeded 0.8 (i.e., 80% of the global maximum). Localisation performance was evaluated by comparing reported and predicted pneumonia-positive lung zones according to precision, sensitivity, and F1-score. Finally, predicted localisations were used as input to generate free-text summaries analogous to the “Impression” section of radiology reports using `DeepSeek-R1-Distill-Llama-8B` (prompt available on GitHub).

## Bibliography

1. GBD 2021 Lower Respiratory Infections and Antimicrobial Resistance Collaborators. Global, regional, and national incidence and mortality burden of non-covid-19 lower respiratory infections and aetiologies, 1990-2021: a systematic analysis from the global burden of disease study 2021. Lancet Infect Dis, 24(9):974–1002, 2024. ISSN 1474-4457 (Electronic) 1473-3099 (Print) 1473-3099 (Linking). doi: 10.1016/S1473-3099(24)00176-2.

2. L. M. Prevedello, S. S. Halabi, G. Shih, C. C. Wu, M. D. Kohli, F. H. Chokshi, B. J. Erickson, J. Kalpathy-Cramer, K. P. Andriole, and A. E. Flanders. Challenges related to artificial intelligence research in medical imaging and the importance of image analysis competitions. Radiol Artif Intell, 1(1):e180031, 2019. ISSN 2638-6100 (Electronic) 2638-6100 (Linking). doi: 10.1148/ryai.2019180031. Prevedello, Luciano M Halabi, Safwan S Shih, George Wu, Carol C Kohli, Marc D Chokshi, Falgun H Erickson, Bradley J Kalpathy-Cramer, Jayashree Andriole, Katherine P Flanders, Adam E eng U01 CA154601/CA/NCI NIH HHS/ U24 CA180918/CA/NCI NIH HHS/ U24 CA180927/CA/NCI NIH HHS/ 2019/01/30 Radiol Artif Intell. 2019 Jan 30;1(1):e180031. doi: 10.1148/ryai.2019180031. eCollection 2019 Jan.

3. A. Makhnevich, L. Sinvani, S. L. Cohen, K. H. Feldhamer, M. Zhang, M. L. Lesser, and T. G. McGinn. The clinical utility of chest radiography for identifying pneumonia: Accounting for diagnostic uncertainty in radiology reports. AJR Am J Roentgenol, 213(6):1207–1212, 2019. ISSN 1546-3141 (Electronic) 0361-803X (Linking). doi: 10.2214/AJR.19.21521. Makhnevich, Alexander Sinvani, Liron Cohen, Stuart L Feldhamer, Kenneth H Zhang, Meng Lesser, Martin L McGinn, Thomas G eng Multicenter Study 2019/09/12 AJR Am J Roentgenol. 2019 Dec;213(6):1207–1212. doi: 10.2214/AJR.19.21521. Epub 2019 Sep 11.

4. M. I. Neuman, E. Y. Lee, S. Bixby, S. Diperna, J. Hellinger, R. Markowitz, S. Servaes, M. C. Monuteaux, and S. S. Shah. Variability in the interpretation of chest radiographs for the diagnosis of pneumonia in children. J Hosp Med, 7(4):294–8, 2012. ISSN 1553-5606 (Electronic) 1553-5592 (Linking). doi: 10.1002/jhm.955. Neuman, Mark I Lee, Edward Y Bixby, Sarah Diperna, Stephanie Hellinger, Jeffrey Markowitz, Richard Servaes, Sabah Monuteaux, Michael C Shah, Samir S eng K01 AI73729/AI/NIAID NIH HHS/ Comparative Study Research Support, N.I.H., Extramural Research Support, Non-U.S. Gov’t 2011/10/20 J Hosp Med. 2012 Apr;7(4):294–8. doi: 10.1002/jhm.955. Epub 2011 Oct 18.

5. G. J. Williams, P. Macaskill, M. Kerr, D. A. Fitzgerald, D. Isaacs, M. Codarini, M. McCaskill, K. Prelog, and J. C. Craig. Variability and accuracy in interpretation of consolidation on chest radiography for diagnosing pneumonia in children under 5 years of age. Pediatr Pulmonol, 48(12):1195–200, 2013. ISSN 1099-0496 (Electronic) 1099-0496 (Linking). doi: 10.1002/ppul.22806. Williams, Gabrielle J Macaskill, Petra Kerr, Marianne Fitzgerald, Dominic A Isaacs, David Codarini, Miriam McCaskill, Mary Prelog, Kristina Craig, Jonathan C eng Research Support, Non-U.S. Gov’t 2013/09/03 Pediatr Pulmonol. 2013 Dec;48(12):1195–200. doi: 10.1002/ppul.22806. Epub 2013 Sep 2.

6. H. G. Chotas and C. E. Ravin. Chest radiography: estimated lung volume and projected area obscured by the heart, mediastinum, and diaphragm. Radiology, 193(2):403–4, 1994. ISSN 0033-8419 (Print) 0033-8419 (Linking). doi: 10.1148/radiology.193.2.7972752. Chotas, H G Ravin, C E eng 1994/11/01 Radiology. 1994 Nov;193(2):403–4. doi: 10.1148/radiology.193.2.7972752.

7. B. Zheng, S. Andrei, M. K. Sarker, and K. D. Gupta. Data Driven Approaches on Medical Imaging. Springer Cham, 1 edition, 2024. ISBN 978-3-031-47772-0. doi: 10.1007/978-3-031-47772-0.

8. Y. C. Peng, W. J. Lee, Y. C. Chang, W. P. Chan, and S. J. Chen. Radiologist burnout: Trends in medical imaging utilization under the national health insurance system with the universal code bundling strategy in an academic tertiary medical centre. Eur J Radiol, 157:110596, 2022. ISSN 1872-7727 (Electronic) 0720-048X (Linking). doi: 10.1016/j.ejrad.2022.110596. Peng, Yan-Chih Lee, Wen-Jeng Chang, Yeun-Chung Chan, Wing P Chen, Shyh-Jye eng Ireland 2022/11/16 Eur J Radiol. 2022 Dec;157:110596. doi: 10.1016/j.ejrad.2022.110596. Epub 2022 Nov 10.

9. P. Rajpurkar, J. Irvin, R. L. Ball, K. Zhu, B. Yang, H. Mehta, T. Duan, D. Ding, A. Bagul, C. P. Langlotz, B. N. Patel, K. W. Yeom, K. Shpanskaya, F. G. Blankenberg, J. Seekins, T. J. Amrhein, D. A. Mong, S. S. Halabi, E. J. Zucker, A. Y. Ng, and M. P. Lungren. Deep learning for chest radiograph diagnosis: A retrospective comparison of the chexnext algorithm to practicing radiologists. PLoS Med, 15(11):e1002686, 2018. ISSN 1549-1676 (Electronic) 1549-1277 (Print) 1549-1277 (Linking). doi: 10.1371/journal.pmed.1002686. Rajpurkar, Pranav Irvin, Jeremy Ball, Robyn L Zhu, Kaylie Yang, Brandon Mehta, Hershel Duan, Tony Ding, Daisy Bagul, Aarti Langlotz, Curtis P Patel, Bhavik N Yeom, Kristen W Shpanskaya, Katie Blankenberg, Francis G Seekins, Jayne Amrhein, Timothy J Mong, David A Halabi, Safwan S Zucker, Evan J Ng, Andrew Y Lungren, Matthew P eng R01 EB000898/EB/NIBIB NIH HHS/ Comparative Study Research Support, Non-U.S. Gov’t Validation Study 2018/11/21 PLoS Med. 2018 Nov 20;15(11):e1002686. doi: 10.1371/journal.pmed.1002686. eCollection 2018 Nov.

10. J. Hofmeister, N. Garin, X. Montet, M. Scheffler, A. Platon, P. A. Poletti, J. Stirnemann, M. P. Debray, Y. E. Claessens, X. Duval, and V. Prendki. Validating the accuracy of deep learning for the diagnosis of pneumonia on chest x-ray against a robust multimodal reference diagnosis: a post hoc analysis of two prospective studies. Eur Radiol Exp, 8(1):20, 2024. ISSN 2509-9280 (Electronic) 2509-9280 (Linking). doi: 10.1186/s41747-023-00416-y. Hofmeister, Jeremy Garin, Nicolas Montet, Xavier Scheffler, Max Platon, Alexandra Poletti, Pierre-Alexandre Stirnemann, Jerome Debray, Marie-Pierre Claessens, Yann-Erick Duval, Xavier Prendki, Virginie eng PRD 7-2015-II/Research and Development Grant of the Geneva University Hospital/ PRD 11-2017-II/Research and Development Grant of the Geneva University Hospital/ England 2024/02/02 Eur Radiol Exp. 2024 Feb 2;8(1):20. doi: 10.1186/s41747-023-00416-y.

11. T. Zhu, K. Xu, W. Son, K. Linton-Reid, M. Boubnovski-Martell, M. Grech-Sollars, A. D. Lain, and J. M. Posma. Designing a computer-assisted diagnosis system for cardiomegaly detection and radiology report generation. PLOS Digit Health, 4(5):e0000835, 2025. ISSN 2767-3170 (Electronic) 2767-3170 (Linking). doi: 10.1371/journal.pdig.0000835. Zhu, Tianhao Xu, Kexin Son, Wonchan Linton-Reid, Kristofer Boubnovski-Martell, Marc Grech-Sollars, Matt Lain, Antoine D Posma, Joram M eng 2025/05/20 PLOS Digit Health. 2025 May 20;4(5):e0000835. doi: 10.1371/journal.pdig.0000835. eCollection 2025 May.

12. B. Allen, S. Agarwal, L. Coombs, C. Wald, and K. Dreyer. 2020 acr data science institute artificial intelligence survey. J Am Coll Radiol, 18(8):1153–1159, 2021. ISSN 1558-349X (Electronic) 1546-1440 (Linking). doi: 10.1016/j.jacr.2021.04.002. Allen, Bibb Agarwal, Sheela Coombs, Laura Wald, Christoph Dreyer, Keith eng 2021/04/24 J Am Coll Radiol. 2021 Aug;18(8):1153–1159. doi: 10.1016/j.jacr.2021.04.002. Epub 2021 Apr 20.

13. R. Wang, P. Chaudhari, and C. Davatzikos. Embracing the disharmony in medical imaging: A simple and effective framework for domain adaptation. Med Image Anal, 76:102309, 2022. ISSN 1361-8423 (Electronic) 1361-8415 (Print) 1361-8415 (Linking). doi: 10.1016/j.media.2021.102309. Wang, Rongguang Chaudhari, Pratik Davatzikos, Christos eng R01 MH112070/MH/NIMH NIH HHS/ RF1 AG054409/AG/NIA NIH HHS/ S10 OD023495/OD/NIH HHS/ U01 AG068057/AG/NIA NIH HHS/ Research Support, N.I.H., Extramural Netherlands 2021/12/07 Med Image Anal. 2022 Feb;76:102309. doi: 10.1016/j.media.2021.102309. Epub 2021 Nov 26.

14. L. Oakden-Rayner. Exploring large-scale public medical image datasets. Acad Radiol, 27 (1):106–112, 2020. ISSN 1878-4046 (Electronic) 1076-6332 (Linking). doi: 10.1016/j.acra.2019.10.006. Oakden-Rayner, Luke eng 2019/11/11 Acad Radiol. 2020 Jan;27(1):106–112. doi: 10.1016/j.acra.2019.10.006. Epub 2019 Nov 6.

15. Ahmed Marey, Parisa Arjmand, Ameerh Dana Sabe Alerab, Mohammad Javad Eslami, Abdelrahman M. Saad, Nicole Sanchez, and Muhammad Umair. Explainability, transparency and black box challenges of ai in radiology: impact on patient care in cardiovascular radiology. Egyptian Journal of Radiology and Nuclear Medicine, 55(1):183, 2024. ISSN 2090-4762. doi: 10.1186/s43055-024-01356-2.

16. A. E. W. Johnson, T. J. Pollard, S. J. Berkowitz, N. R. Greenbaum, M. P. Lungren, C. Y. Deng, R. G. Mark, and S. Horng. Mimic-cxr, a de-identified publicly available database of chest radiographs with free-text reports. Sci Data, 6(1):317, 2019. ISSN 2052-4463 (Electronic) 2052-4463 (Linking). doi: 10.1038/s41597-019-0322-0. Johnson, Alistair E W Pollard, Tom J Berkowitz, Seth J Greenbaum, Nathaniel R Lungren, Matthew P Deng, Chih-Ying Mark, Roger G Horng, Steven eng R01 EB017205/EB/NIBIB NIH HHS/ R01 GM104987/GM/NIGMS NIH HHS/ NIH-R01-EB017205/U.S. Department of Health amp; Human Services | National Institutes of Health (NIH)/International Dataset Research Support, N.I.H., Extramural England 2019/12/14 Sci Data. 2019 Dec 12;6(1):317. doi: 10.1038/s41597-019-0322-0.

17. Jeremy Irvin, Pranav Rajpurkar, Michael Ko, Yifan Yu, Silviana Ciurea-Ilcus, Chris Chute, Henrik Marklund, Behzad Haghgoo, Robyn Ball, Katie Shpanskaya, Jayne Seekins, David A. Mong, Safwan S. Halabi, Jesse K. Sandberg, Ricky Jones, David B. Larson, Curtis P. Langlotz, Bhavik N. Patel, Matthew P. Lungren, and Andrew Y. Ng. Chexpert: A large chest radiograph dataset with uncertainty labels and expert comparison. Proceedings of the AAAI Conference on Artificial Intelligence, 33(01):590–597, 2019. doi: 10.1609/aaai.v33i01.3301590.

18. A. Bustos, A. Pertusa, J. M. Salinas, and M. de la Iglesia-Vaya. Padchest: A large chest x-ray image dataset with multi-label annotated reports. Med Image Anal, 66:101797, 2020. ISSN 1361-8423 (Electronic) 1361-8415 (Linking). doi: 10.1016/j.media.2020.101797. Bustos, Aurelia Pertusa, Antonio Salinas, Jose-Maria de la Iglesia-Vaya, Maria eng Research Support, Non-U.S. Gov’t Netherlands 2020/09/03 Med Image Anal. 2020 Dec;66:101797. doi: 10.1016/j.media.2020.101797. Epub 2020 Aug 20.

19. Xiaosong Wang, Yifan Peng, L. Lu, Zhiyong Lu, Mohammadhadi Bagheri, and Ronald M. Summers. Chestx-ray8: Hospital-scale chest x-ray database and benchmarks on weakly-supervised classification and localization of common thorax diseases, 2017.

20. G. Shih, C. C. Wu, S. S. Halabi, M. D. Kohli, L. M. Prevedello, T. S. Cook, A. Sharma, J. K. Amorosa, V. Arteaga, M. Galperin-Aizenberg, R. R. Gill, M. C. B. Godoy, S. Hobbs, J. Jeudy, Laroia, P. N. Shah, D. Vummidi, K. Yaddanapudi, and A. Stein. Augmenting the national institutes of health chest radiograph dataset with expert annotations of possible pneumonia. Radiol Artif Intell, 1(1):e180041, 2019. ISSN 2638-6100 (Electronic) 2638-6100 (Linking). doi: 10.1148/ryai.2019180041. Shih, George Wu, Carol C Halabi, Safwan S Kohli, Marc D Prevedello, Luciano M Cook, Tessa S Sharma, Arjun Amorosa, Judith K Arteaga, Veronica Galperin-Aizenberg, Maya Gill, Ritu R Godoy, Myrna C B Hobbs, Stephen Jeudy, Jean Laroia, Archana Shah, Palmi N Vummidi, Dharshan Yaddanapudi, Kavitha Stein, Anouk eng 2019/01/30 Radiol Artif Intell. 2019 Jan 30;1(1):e180041. doi: 10.1148/ryai.2019180041. eCollection 2019 Jan.

21. H. Q. Nguyen, K. Lam, L. T. Le, H. H. Pham, D. Q. Tran, D. B. Nguyen, D. D. Le, C. M. Pham, H. T. T. Tong, D. H. Dinh, C. D. Do, L. T. Doan, C. N. Nguyen, B. T. Nguyen, Q. V. Nguyen, A. D. Hoang, H. N. Phan, A. T. Nguyen, P. H. Ho, D. T. Ngo, N. T. Nguyen, N. T. Nguyen, M. Dao, and V. Vu. Vindr-cxr: An open dataset of chest x-rays with radiologist’s annotations. Sci Data, 9(1):429, 2022. ISSN 2052-4463 (Electronic) 2052-4463 (Linking). doi: 10.1038/s41597-022-01498-w. Nguyen, Ha Q Lam, Khanh Le, Linh T Pham, Hieu H Tran, Dat Q Nguyen, Dung B Le, Dung D Pham, Chi M Tong, Hang T T Dinh, Diep H Do, Cuong D Doan, Luu T Nguyen, Cuong N Nguyen, Binh T Nguyen, Que V Hoang, Au D Phan, Hien N Nguyen, Anh T Ho, Phuong H Ngo, Dat T Nguyen, Nghia T Nguyen, Nhan T Dao, Minh Vu, Van eng Dataset England 2022/07/21 Sci Data. 2022 Jul 20;9(1):429. doi: 10.1038/s41597-022-01498-w.

22. S. M. Pizer, E. P. Amburn, J. D. Austin, R. Cromartie, A. Geselowitz, T. Greer, B. Terhaarromeny, J. B. Zimmerman, and K. Zuiderveld. Adaptive histogram equalization and its variations. Computer Vision Graphics and Image Processing, 39(3):355–368, 1987. ISSN 0734-189x. doi: Doi 10.1016/S0734-189x(87)80186-X. J5672 Times Cited:2534 Cited References Count:10.

23. G. Huang, Z. Liu, L. Van Der Maaten, and K. Q. Weinberger. Densely connected convolutional networks. In 2017 IEEE Conference on Computer Vision and Pattern Recognition (CVPR), pages 2261–2269, 2017. ISBN 1063-6919. doi: 10.1109/CVPR.2017.243.

24. R. R. Selvaraju, M. Cogswell, A. Das, R. Vedantam, D. Parikh, and D. Batra. Grad-cam: Visual explanations from deep networks via gradient-based localization. In 2017 IEEE International Conference on Computer Vision (ICCV), pages 618–626, 2017. ISBN 2380-7504. doi: 10.1109/ICCV.2017.74.

25. N. Otsu. A threshold selection method from gray-level histograms. IEEE Transactions on Systems, Man, and Cybernetics, 9(1):62–66, 1979. ISSN 2168-2909. doi: 10.1109/TSMC.1979.4310076.

26. Sarah Sandmann, Stefan Hegselmann, Michael Fujarski, Lucas Bickmann, Benjamin Wild, Roland Eils, and Julian Varghese. Benchmark evaluation of deepseek large language models in clinical decision-making. Nature Medicine, 2025. ISSN 1546-170X. doi: 10.1038/s41591-025-03727-2.

27. A. Abdullah and S. T. Kim. Automated radiology report labeling in chest x-ray pathologies: Development and evaluation of a large language model framework. JMIR Med Inform, 13: e68618, 2025. ISSN 2291-9694 (Electronic) 2291-9694 (Linking). doi: 10.2196/68618. Abdullah, Abdullah Kim, Seong Tae eng Canada 2025/03/28 JMIR Med Inform. 2025 Mar 28;13:e68618. doi: 10.2196/68618.

28. Francesco Corradi, Claudia Brusasco, Alessandro Garlaschi, Francesco Paparo, Lorenzo Ball, Gregorio Santori, Paolo Pelosi, Fiorella Altomonte, Antonella Vezzani, and Vito Brusasco. Quantitative analysis of lung ultrasonography for the detection of communityacquired pneumonia: A pilot study. BioMed Research International, 2015(1):868707, 2015. ISSN 2314-6133. doi: 10.1155/2015/868707.

29. Xiao-lei Liu, Rui Lian, Yong-kang Tao, Cheng-dong Gu, and Guo-qiang Zhang. Lung ultrasonography: an effective way to diagnose community-acquired pneumonia. Emergency Medicine Journal, 32(6):433, 2015. doi: 10.1136/emermed-2013-203039.

30. J. R. Zech, M. A. Badgeley, M. Liu, A. B. Costa, J. J. Titano, and E. K. Oermann. Variable generalization performance of a deep learning model to detect pneumonia in chest radiographs: A cross-sectional study. PLoS Med, 15(11):e1002683, 2018. ISSN 1549-1676 (Electronic) 1549-1277 (Print) 1549-1277 (Linking). doi: 10.1371/journal.pmed.1002683. Zech, John R Badgeley, Marcus A Liu, Manway Costa, Anthony B Titano, Joseph J Oermann, Eric Karl eng Multicenter Study Research Support, Non-U.S. Gov’t 2018/11/07 PLoS Med. 2018 Nov 6;15(11):e1002683. doi: 10.1371/journal.pmed.1002683. eCollection 2018 Nov.

31. Brad H. Thompson, Kevin S. Berbaum, Michael J. George, and John W. Ely. Identifying left lower lobe pneumonia at chest radiography: Performance of family practice residents before and after a didactic session. Academic Radiology, 5(5):324–328, 1998. ISSN 1076-6332. doi: 10.1016/S1076-6332(98)80150-5.

32. Harsh Panwar, P. K. Gupta, Mohammad Khubeb Siddiqui, Ruben Morales-Menendez, Prakhar Bhardwaj, and Vaishnavi Singh. A deep learning and grad-cam based color visualization approach for fast detection of covid-19 cases using chest x-ray and ct-scan images. Chaos, Solitons Fractals, 140:110190, 2020. ISSN 0960-0779. doi: 10.1016/j.chaos.2020.110190.

33. Aki Miyazaki, Kengo Ikejima, Mizuho Nishio, Minoru Yabuta, Hidetoshi Matsuo, Koji Onoue, Takaaki Matsunaga, Eiko Nishioka, Atsushi Kono, Daisuke Yamada, Ken Oba, Reiichi Ishikura, and Takamichi Murakami. Computer-aided diagnosis of chest x-ray for covid-19 diagnosis in external validation study by radiologists with and without deep learning system. Scientific Reports, 13(1):17533, 2023. ISSN 2045-2322. doi: 10.1038/s41598-023-44818-9.

